# ASSOCIATION BETWEEN CIRCADIAN RHYTHMS, SLEEP, AND DEPRESSION: A BIDIRECTIONAL MENDELIAN RANDOMISATION STUDY IN THE UK BIOBANK

**DOI:** 10.1101/2024.12.08.24318680

**Authors:** Valentina Paz, Dylan M Williams, Marcus Richards, Bettina Tassino, Ana Silva, Victoria Garfield

## Abstract

Increasing evidence links circadian and sleep disruptions with depression, though whether they are causally related remains unclear. This study used Mendelian randomization (MR) with exposures instrumented using genome-wide genetic variants and data from the UK Biobank (n=408 480; mean age=56.9 years) to explore causal associations between circadian and sleep traits, and depression. We conducted bidirectional MR analyses between morningness, sleep duration, insomnia symptoms, and depression, and secondary unidirectional analyses with circadian and sleep traits as exposures and mixed anxiety-depressive symptoms and anxiety disorders as outcomes. Using inverse-variance weighting and sensitivity analyses for horizontal pleiotropy (MR-Egger and Weighted Median Estimator), we found suggestive associations between morningness and lower odds of depression (diagnosis and treatment: OR=0.89, 95%CI=0.80, 0.98), and depression and shorter sleep duration (β=-4.67 minutes, 95%CI=-7.44; -1.89). Depression showed an effect on insomnia (any symptoms: OR=1.22, 95%CI=1.11; 1.34; frequent symptoms: OR=1.30, 95%CI=1.19; 1.42), while frequent insomnia was associated with increased odds of depression (diagnosis: OR=1.14, 95%CI=1.10; 1.19; diagnosis and treatment: OR=1.10, 95%CI=1.06; 1.14). Insomnia also had an effect on mixed anxiety-depressive symptoms (any symptoms: OR=1.19, 95%CI=1.08, 1.32; frequent symptoms: OR=1.20, 95%CI=1.17, 1.23), but no associations were found between the exposures and anxiety disorders. These findings shed light on the causal links between the circadian system and depression.

## Introduction

The circadian system controls the endogenous rhythmic activity of numerous physiological functions in humans, the sleep-wake cycle being the most evident circadian rhythm [1]. Sleep is a complex behaviour characterised by multiple dimensions. Some of the most important dimensions of sleep are timing, the placement of sleep within the 24-hour day; duration, defined as the total amount of sleep obtained over 24 hours [2]; and quality, understood as the self-satisfaction with sleep [3]. Circadian disruption, considered the consequence of a temporal challenge or other perturbations in the circadian system [4], is a critical factor in numerous circadian and sleep disorders and has been linked to several adverse health outcomes, including cardiovascular, metabolic, and mental disorders [5].

Depression, a prevalent mental disorder and the leading cause of years of life lost due to disability worldwide, is characterized by persistent low mood, anhedonia, and high comorbidity with anxiety disorders [6]. This disorder is often associated with circadian and sleep disturbances [7–9]. These disturbances usually persist across different phases of the disorder and contribute to increased treatment resistance [8]. Moreover, sleep problems are recognized as a vegetative sign of depression [10] and one of its most common residual symptoms [11]. The association between depression and circadian preferences, the preference to time daily activities including sleep used as an estimate of circadian phase, has been extensively reported in cross-sectional studies, which indicate that evening types are more prone to depression [12–14]. Epidemiological research further suggests that both short and long sleep durations are associated with depression [15], and short sleep often persists even after the remission of emotional symptoms [16]. Insomnia —characterized by disturbances in sleep continuity, including early morning awakenings, difficulty falling asleep and staying asleep, and non-restorative or poor-quality sleep— has been historically considered an epiphenomenon of depression. However, recent longitudinal studies suggest that insomnia is a risk factor for the development of the disorder [17,18]. However, the causal nature of these associations remains elusive [19].

Mendelian randomisation (MR), which uses genetic variants to help assess causality, could be used to address these questions [20]. Regarding the association between circadian preferences and depression, some studies found that the risk of depression is lower in morning individuals [21,22]. Moreover, recent work has revealed that short sleep duration has an effect on the risk of depression [23] and bidirectional associations between sleep duration and depression using MR [24]. Studies also reported evidence of an effect of insomnia symptoms on depressive symptoms [25] and risk of depression [23,26–28] and bidirectional associations between insomnia and depression [29–33]. However, other studies have not found causal associations between circadian and sleep traits and depression, such as duration [34], morningness [23,35] or insomnia [36].

Considering that this field of study is still in development and that results reported to date are ambiguous, it is crucial to investigate whether the association between circadian preferences and sleep patterns and depression might be causal, and if so, to determine the directionality of this association. Furthermore, considering the comorbidity between depression and anxiety [6], this study aims to elucidate the role of anxiety in these associations. Specifically, the present study examines whether circadian and sleep phenotypes are causally associated with depression.

## Materials and Methods

### Sample

Study participants were from the UK Biobank (UKB) cohort, described in detail elsewhere [37]. The UKB is a large-scale biomedical database containing phenotypic and genetic data available for over 500 000 participants from the general United Kingdom population aged between 40 and 69 at recruitment (2006 to 2010). We used data on up to 408 480 white British individuals (genetically defined) from the UKB with quality-controlled genotype and phenotype data available. The mean age of participants was 56.9 ± 8.0 years, 54.0% were female, and 19.9% were in the most deprived socioeconomic quintile (Table 2 for full sample characteristics).

**Table 1.**
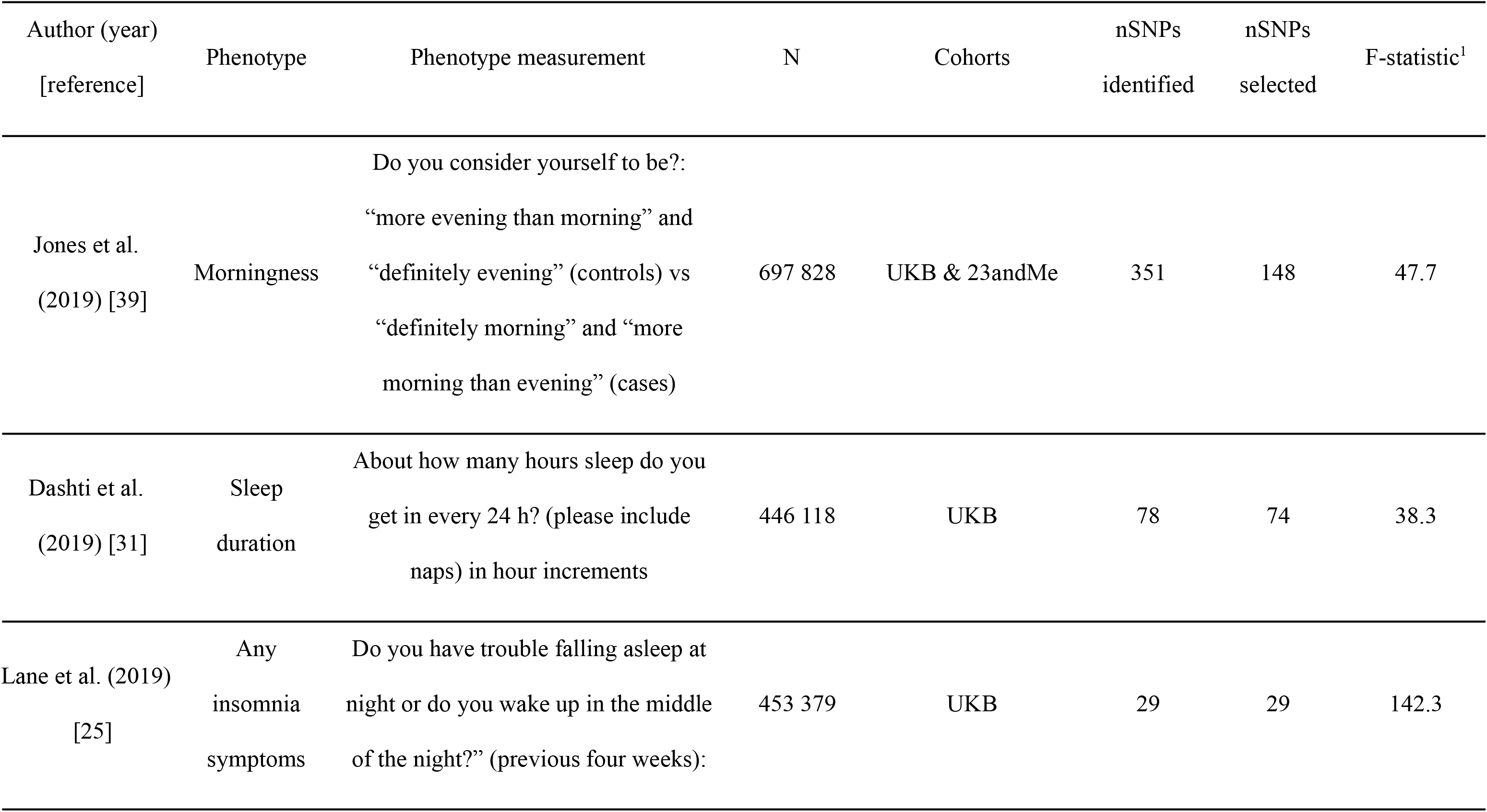
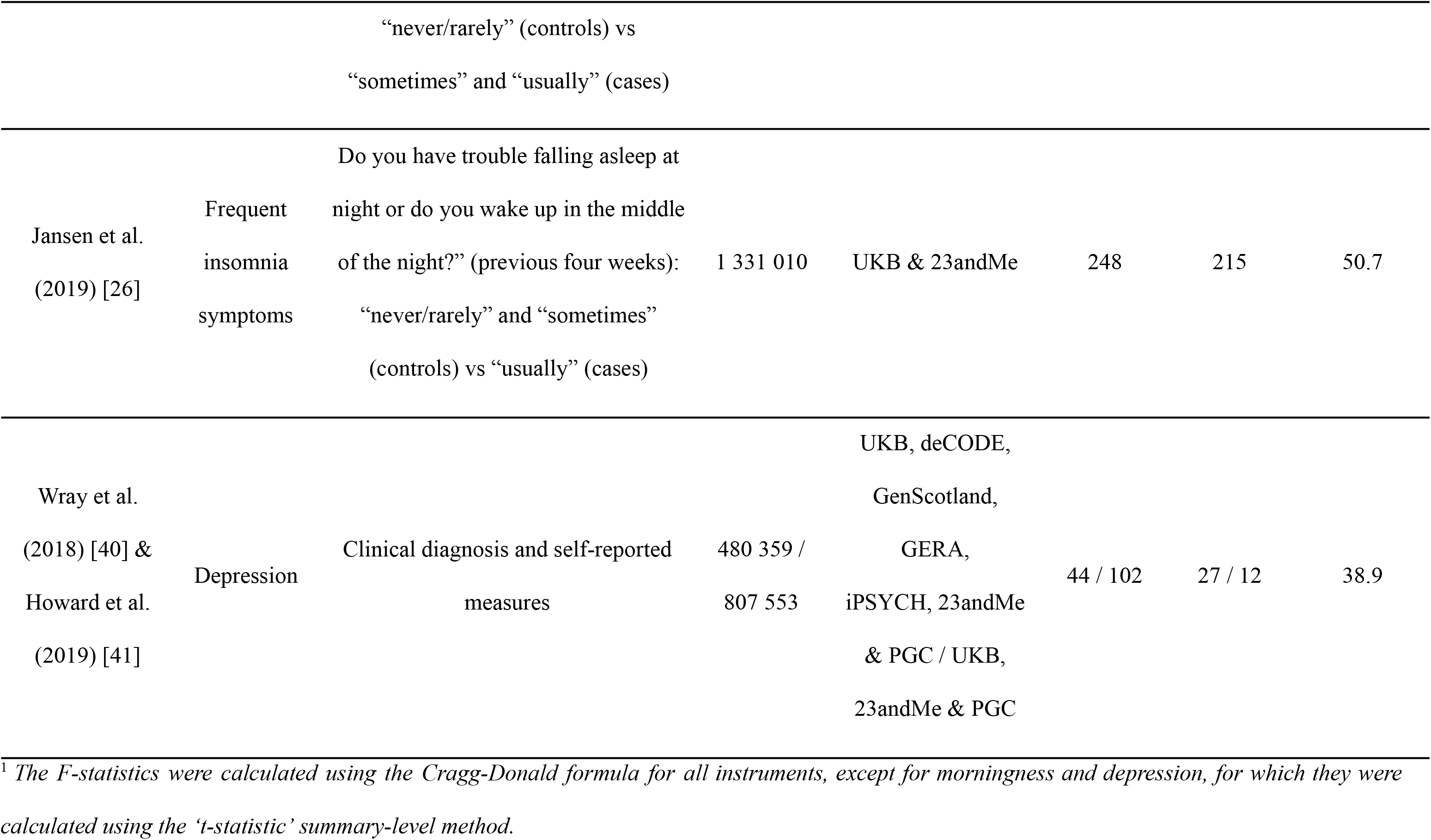
Summary of genetic instruments.

**Table 2.**
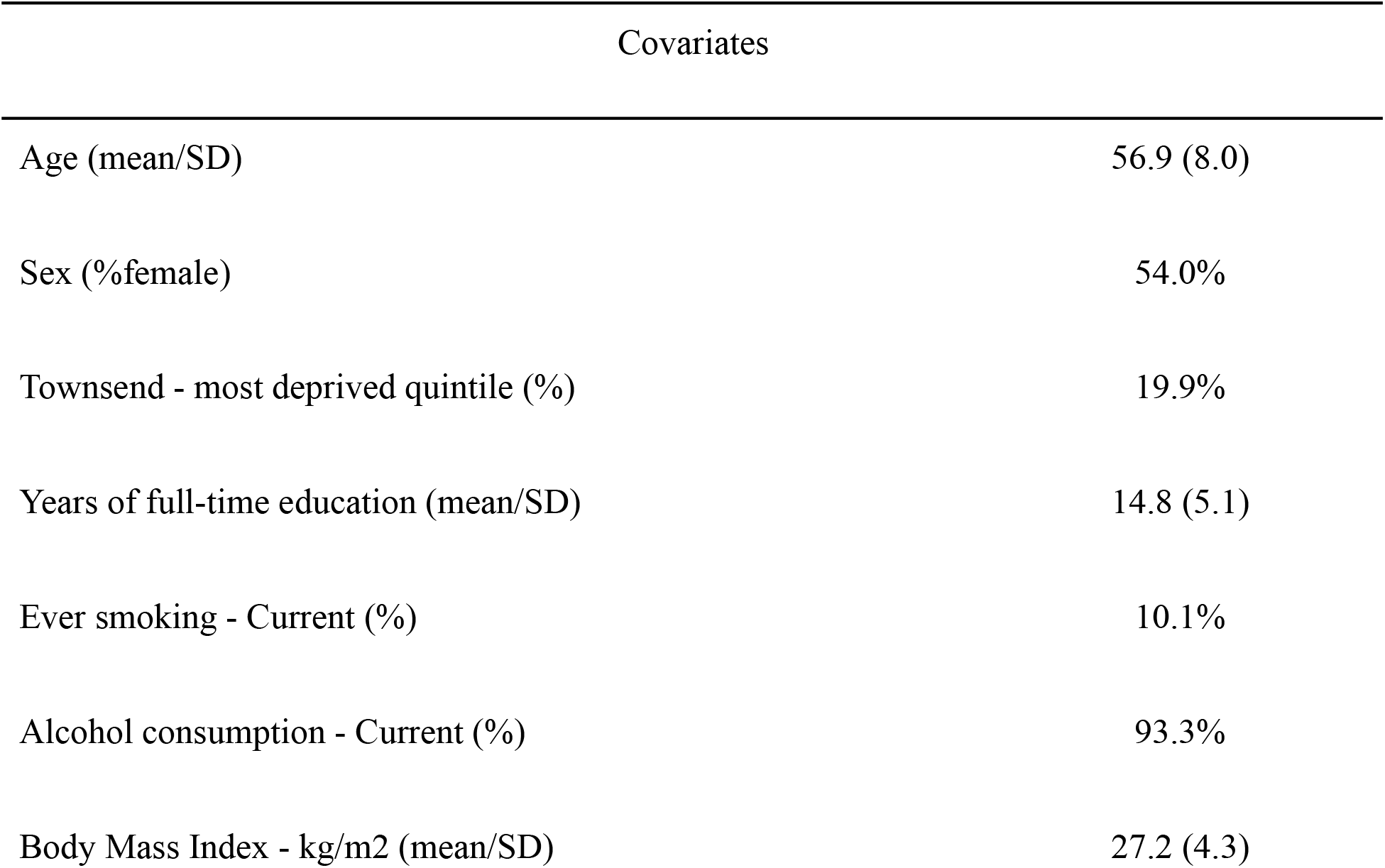
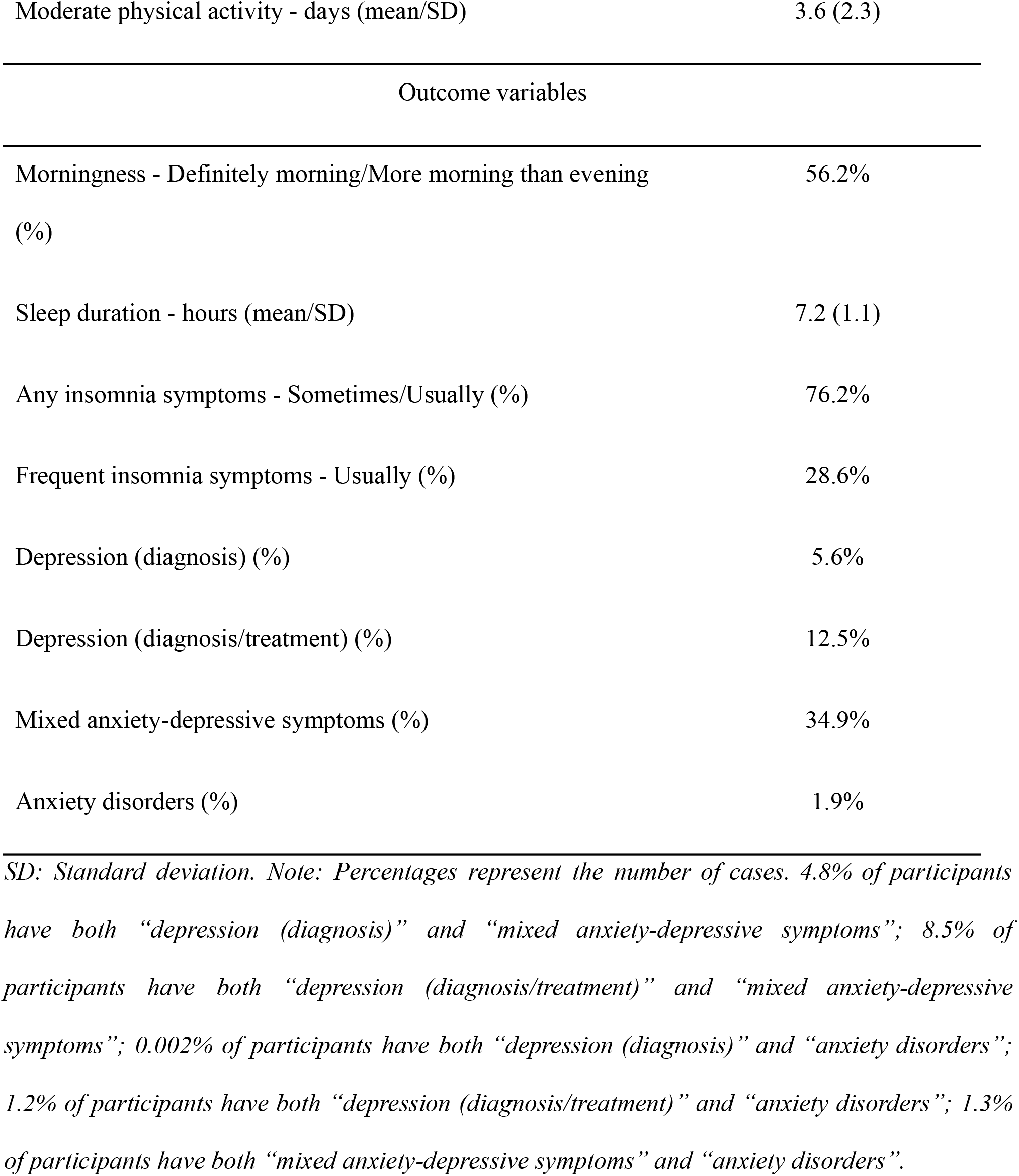
Sample characteristics.

| Covariates |  |
| --- | --- |
| Age (mean/SD) | 56.9 (8.0) |
| Sex (%female) | 54.0% |
| Townsend - most deprived quintile (%) | 19.9% |
| Years of full-time education (mean/SD) | 14.8 (5.1) |
| Ever smoking - Current (%) | 10.1% |
| Alcohol consumption - Current (%) | 93.3% |
| Body Mass Index - kg/m<sup>2</sup> (mean/SD) | 27.2 (4.3) |
| Moderate physical activity - days (mean/SD) | 3.6 (2.3) |
| Outcome variables |  |
| Morningness - Definitely morning/More morning than evening (%) | 56.2% |
| Sleep duration - hours (mean/SD) | 7.2 (1.1) |
| Any insomnia symptoms - Sometimes/Usually (%) | 76.2% |
| Frequent insomnia symptoms - Usually (%) | 28.6% |
| Depression (diagnosis) (%) | 5.6% |
| Depression (diagnosis/treatment) (%) | 12.5% |
| Mixed anxiety-depressive symptoms (%) | 34.9% |
| Anxiety disorders (%) | 1.9% |

*SD: Standard deviation. Note: Percentages represent the number of cases. 4.8% of participants have both “depression (diagnosis)” and “mixed anxiety-depressive symptoms”; 8.5% of participants have both “depression (diagnosis/treatment)” and “mixed anxiety-depressive symptoms”; 0.002% of participants have both “depression (diagnosis)” and “anxiety disorders”; 1.2% of participants have both “depression (diagnosis/treatment)” and “anxiety disorders”; 1.3% of participants have both “mixed anxiety-depressive symptoms” and “anxiety disorders”.*

### Study design

This study used a pseudo two-sample MR design to estimate bidirectional associations between circadian and sleep traits, and depression and anxiety. Thus, we used a combination of genetic association estimates that we derived from individual-level UKB data and GWAS summary statistics, some of which analysed only UKB data, and others that analysed UKB alongside other datasets. This study followed the STROBE-MR reporting guidelines [38].

### Genotyping and quality control

487 409 UKB participants were genotyped using one of two customised genome-wide arrays imputed to a combination of the UK10K, 1000 Genomes Phase 3, and the Haplotype Reference Consortium (HRC) reference panels, which resulted in data on 93 095 623 autosomal variants [39]. We then applied additional variant-level quality control (QC) and excluded genetic variants with Fisher’s exact test <0.3, minor allele frequency (MAF) <1%, and a missing call rate of ≥5%.

Participants were excluded if they had excessive or minimal heterozygosity, more than 10 putative third-degree relatives as per the kinship matrix, no consent to extract DNA, sex mismatches between self-reported and genetic sex, missing QC information and non-European ancestry (based on self-reported ancestry and the similarity with their “genetic ancestry” [40], as per a principal component analysis of their genotype) (see Lu et al. 2002 [41] for the differences between race, ethnicity, and ancestry).

### Selection of genetic instruments

Morningness was instrumented using independent genetic variants (at LD cut-off r^2^≤0.3) from a GWAS by Jones et al. (2019) [42]. The variants were discovered in 697 828 European participants using data from the UKB cohort and summary statistics from a GWAS performed with 23andMe data (we used only UKB summary statistics). In the UKB cohort, circadian preferences was assessed based on the question “Do you consider yourself to be?” administered at baseline, with possible responses “More evening than morning” and “Definitely evening” coded as controls and “Definitely morning” and “More morning than evening” coded as cases (“Do not know” and “Prefer not to answer” responses were coded as missing). Of the respondents, 23.9% identified as “Definitely morning”, 32.2% as “More morning than evening”, 25.6% as “More evening than morning”, 8.0% as “Definitely evening”, and 10.3% as “Do not know”. Of the 351 variants discovered, we used 148.

Sleep duration was instrumented using independent genetic variants (at LD cut-off r^2^<0.05 and 1 Mb) discovered in a 2019 GWAS of 446 118 UKB European participants [34], which explained 0.69% of the variance in sleep duration. These variants were associated with the question “About how many hours sleep do you get in every 24 h? (please include naps)” administered at baseline, with responses in hourly increments (authors excluded responses >3H or <18h and, “Do not know” and “Prefer not to answer” responses were coded as missing). The authors reported a mean self-reported habitual sleep duration of 7.2 hours. Of the 78 variants discovered, we used 74.

Any insomnia symptoms were instrumented using 29 independent genetic variants from a previously published large-scale GWAS [25]. These variants were discovered in 453 379 UKB European participants, explaining 1.0% of the variance, based on the question “Do you have trouble falling asleep at night or do you wake up in the middle of the night?” administered at baseline, with possible responses “Never/rarely” defined as controls and “Sometimes” and “Usually” defined as cases (“Prefer not to answer” responses were coded as missing). If the participants asked for help, they were shown the message “if this varies a lot, answer this question in relation to the last 4 weeks”. Of the UKB respondents, 23.9% responded “Never/rarely” and 76.1% “Sometimes” or “Usually”. The authors also performed another GWAS for frequent insomnia symptoms not used here, as we decided to use Jansen et al.’s (2019) GWAS due to how cases and controls were defined [32].

Frequent insomnia symptoms were instrumented using independent genetic variants (at LD cut-off r^2^≤0.6 and >250 kb) discovered in the GWAS of Jansen et al. (2019) [32]. These variants were discovered in 1 331 010 European participants from the UKB and 23andMe cohorts combined and explained 2.6% of the variance. The lead genetic variants showed concordant effects in both samples. For UKB participants, the same question used in the GWAS by Lane et al. (2019) was employed. Responses “Never/rarely” and “Sometimes” were defined as controls and “Usually” as cases. For 23andMe participants, insomnia cases were defined as those who affirmed being told by a doctor, diagnosed or treated for insomnia, diagnosed with sleep disturbances, routinely having trouble getting to sleep at night, or ever taking prescription sleep aids. Controls were defined as those who did not provide a positive or uncertain response (e.g. "I don’t know"; "I am not sure") to any of the issues listed above, nor were diagnosed with Narcolepsy, Sleep apnea, Restless leg syndrome, Post-traumatic stress disorder, Autism, or Asperger. The prevalence of insomnia was 28.3% in the UKB sample, 30.5% in 23andMe, and 29.9% in the combined sample. Of the 248 variants discovered, we used 215.

Depression was instrumented using independent genetic variants discovered in two large-scale GWAS [43,44] to increase the number of valid variants following criteria from our recent review [45]. Wray et al. (2018) [44] conducted a genome-wide meta-analysis of 480 359 Europeans from seven different cohorts (including UKB), explaining 1.9% of the variance (at LD cut-off r^2^<0.1). Cases (28.2%) were defined as those with an MDD diagnosis established using structured diagnostic instruments from assessments by trained interviewers, self-reported symptoms, diagnosis or treatment, or medical records. Controls (71.8%) were screened for the absence of lifetime Major Depressive Disorder (MDD) and randomly selected from the population. Variants from Howard et al. (2019) [43] were discovered in a genome-wide meta-analysis of 807 553 Europeans from three large studies (LD cut-off r^2^<0.1 and 3 Mb). The analysis included the 23andMe_307k cohort [46], the UKB cohort [47], and the PGC_139k cohort [44] (we used only UKB summary statistics). The depression phenotype included probable MDD based on self-reported depressive symptoms with associated impairment, MDD identified from hospital admission records and self-reported help-seeking for problems with nerves, anxiety, tension, or depression. Controls were 69.5% of the total sample, while cases were 30.5%. Of the 146 variants discovered, we used 39 variants.

Following guidance from our recent review of selection of sleep genetic instruments [45], we selected genetic instruments for morningness, sleep duration, and insomnia symptoms (Table 1 and Supplementary Table 1). Genetic instruments for depression were selected following the same criteria. In particular, we excluded correlated variants using linkage disequilibrium (LD) clumping with r2<0.01 within 250kb. We only included genome-wide significant (p<5x10^-8^) variants. We excluded weak instruments, as assessed by the F-statistics (F>10 indicates that substantial weak instrument bias is unlikely [48]) using the Cragg-Donald formula (F=(n−k−1/k) (R^2^/1−R^2^)) [49] (k = number of instruments) or the ‘t-statistic’ summary-level method when the R^2^ was unknown [45]. We also harmonised the genetic variants between the exposure GWAS and our outcome sample by aligning effect alleles. We did not exclude palindromic variants because we mostly used a single sample. Finally, we calculated the potential bias due to overlapping samples. This bias implies overestimating the strongest variant in the data under analysis when using genetic variants discovered in the analytical sample [50]. We used an online calculator available at https://sb452.shinyapps.io/overlap/ [51] assuming 100% sample overlap. We estimated that the biases were small for all outcomes analysed (absolute value of biases ≤0.001) with type-1 error rate = 0.05 (Supplementary Table 2).

### Circadian and sleep measures

We selected our morningness, sleep duration, and insomnia symptoms outcomes using phenotypes similar to our exposures. For morningness, we used the same questions and categories as Jones et al. (2019) [42]. For sleep duration, we used the same question as Dashti et al. (2019) [34] with responses in hourly increments. Based on our sleep duration distribution, we excluded extreme responses of <2H or >12h. For any and frequent insomnia symptoms, we used the same UKB question and categories as Lane et al. (2019) [25] and Jansen et al. (2019) [32], respectively.

### Depression measures

For depression, we pragmatically decided to use two phenotypes: “diagnosis” (primary outcome for depression) and “diagnosis or treatment” (secondary outcome for depression). The diagnosis category was composed of participants who had answered yes to the question “Have you been diagnosed with one or more of the following mental health problems by a professional, even if you don’t have it currently?: depression” plus participants with primary or secondary MDD (mild, moderate, severe or recurrent without psychotic symptoms) identified from hospital inpatient records.

The diagnosis and treatment category was composed of participants in the diagnosis category plus those who have ever taken substances (prescribed medication for at least two weeks, unprescribed medication more than once or drugs or alcohol more than once) or undertaken certain activities more than once (therapies such as psychotherapy, counselling, group therapy or cognitive-behavioural therapy or other therapeutic activities such as mindfulness, yoga or art classes) to cope with symptoms of depression (prolonged feelings of sadness and/or anhedonia). We included this secondary outcome to increase the proportion of cases with the aim of replicating our results. However, this outcome is less precise in assessing depression compared to the primary outcome, as it encompasses a broad spectrum of therapeutic strategies to manage depressive symptoms.

### Anxiety measures

Considering the comorbidity between depression and anxiety [6] and the fact that the genetic variants discovered for depression by Howard et al. (2019) [43] were based on a phenotype that included self-reported help-seeking for problems with nerves, anxiety, tension, or depression, we also analysed two outcomes related to anxiety. We did not conduct bidirectional analyses with anxiety since the SNPs discovered to date have not been replicated at the genome-wide level of significance [52].

The mixed anxiety-depressive symptoms outcome was composed of participants who had answered yes to at least one of the following questions: “Have you ever seen a general practitioner (GP) for nerves, anxiety, tension or depression?” and “Have you ever seen a psychiatrist for nerves, anxiety, tension or depression?”.

The anxiety disorders outcome was composed of participants who had answered yes to the question, “Have you been diagnosed with one or more of the following mental health problems by a professional, even if you don’t have it currently?: “anxiety, nerves or generalised anxiety disorder”. This category also included participants with primary or secondary “Generalised anxiety disorder” identified from hospital inpatient records.

### Statistical analyses

#### i. Main analyses

We performed logistic regression between each of the genetic variants and our outcomes (for sleep duration as an outcome we performed linear regressions), adjusting for ten principal components to minimise issues of residual confounding by population stratification (for the analyses of depression to morningness, insomnia, and sleep duration, we corroborated that when adjusting for anxiety disorders the results remained the same). Inverse-variance weighting (IVW) MR was implemented. The IVW, also known as ‘conventional MR’ estimates the effect of an exposure (e.g., depression) on a given outcome (e.g., insomnia) by taking an average of the genetic variants’ ratio of variant-outcome (SNP→Y) to variant-exposure (SNP→X) association, which is calculated using the principles of a fixed-effects meta-analysis [53]. We also implemented standard sensitivity analyses, including MR-Egger and the Weighted Median Estimator (WME). MR-Egger regression yields an intercept term to denote the presence or absence of unbalanced horizontal pleiotropy and its estimand accounts for this pleiotropy [54], while the WME can give more robust estimates when up to 50% of the genetic variants have invalid weights. Consistency in effect sizes and overlap of 95% confidence intervals (CI) were examined [55]. Results are expressed as odds ratios, which should be interpreted as odds of having the outcome for 1-log-odd of increase in the exposure. Only for sleep duration as an outcome, results are expressed as unstandardised beta coefficients, which should be interpreted as differences in the outcome for every 1-unit increase in the exposure. Analyses were performed using PLINK 2.0 and the “MendelianRandomization” package [56] for R studio version 2022.02.0.

#### ii. Additional analyses

Following the same approach as for our main analyses, we tested the association between our circadian and sleep exposures and mixed anxiety-depressive symptoms, and anxiety disorders as outcomes.

#### iii. Testing of MR assumptions

a. Associations between the genetic instrument and exposure instrumented (GWAS robust): variants instrumented have been robustly associated with our phenotypes in recent large-scale GWAS.

b. No association between genetic instruments and the outcome other than via the exposure under study (no horizontal pleiotropy): we implemented MR-Egger and WME sensitivity analyses to test this assumption.

c. No confounding of the SNP-outcome associations (e.g. confounding by population stratification, assortative mating or dynastic effects): we regressed several covariates on our main instruments using a Bonferroni multiple testing correction of 0.05/nSNPs. The list of covariates we selected was based on recent literature [57–59] and included age, sex (female, male), years of full-time education, deprivation (Townsend deprivation quintiles), smoking status (never or previous vs. current smoker), alcohol status (never or previous vs current consumer), body mass index (BMI) (kg/m2) and, physical activity (days of moderate activity for more than 10 minutes).

## Results

### Main MR results

#### Associations between morningness and depression

No associations were found between genetically predicted morningness and diagnosis of depression (OR=0.96, 95%CI=0.87; 1.06) (Figure 1a, Supplementary Tables 3 & 7). Associations were found between genetically predicted morningness and lower odds of having a diagnosis or treatment of depression only with the WME (OR=0.89, 95%CI=0.80; 0.98) (Figure 1b, Supplementary Tables 3 & 7). No associations were found between genetically predicted depression and morningness (OR=1.01, 95%CI=0.90; 1.14) (Figure 1c, Supplementary Tables 3 & 7).

**Figure 1.**
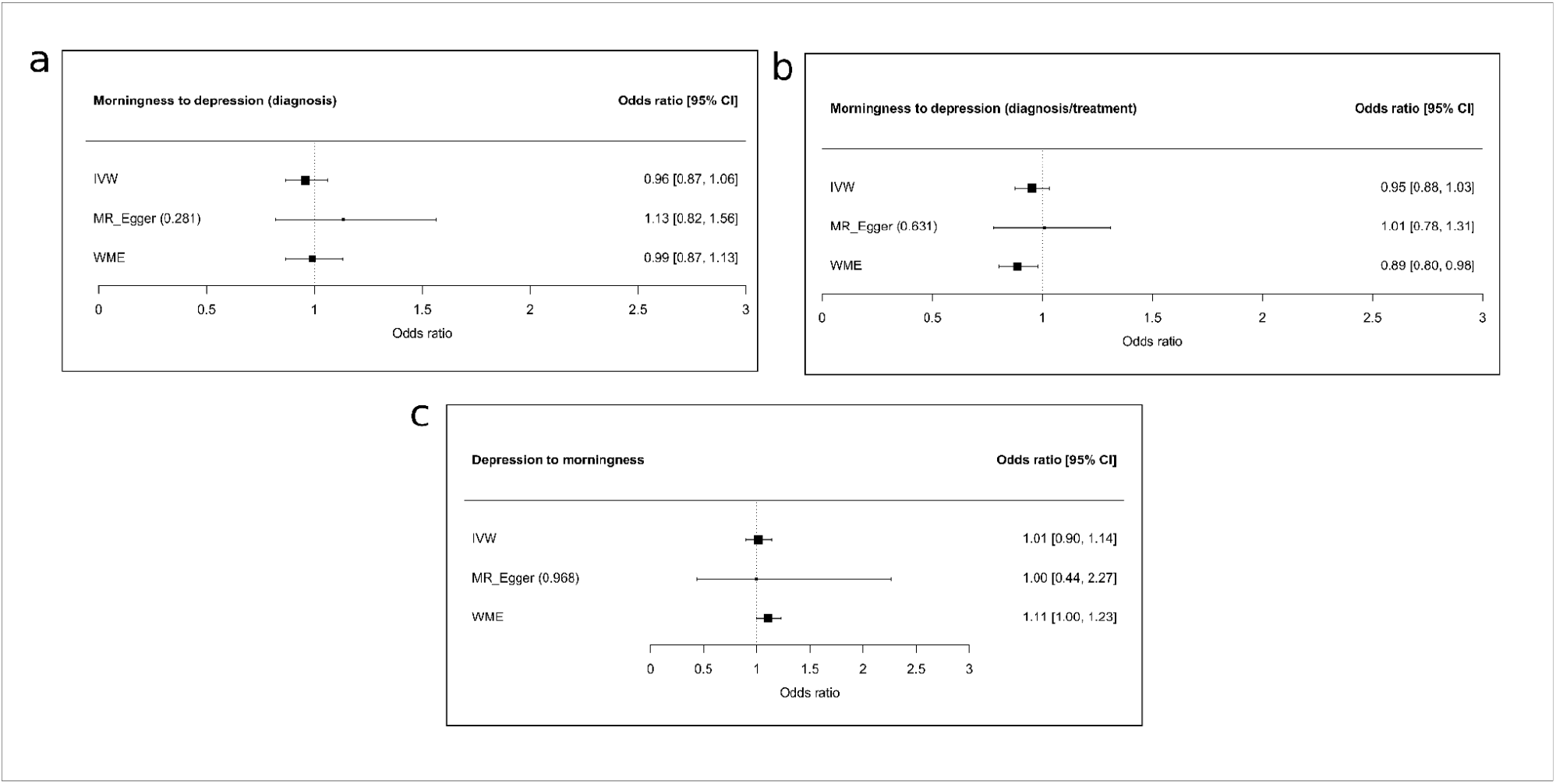
Associations between morningness and depression, including sensitivity analyses. a) Morningness to depression (diagnosis). b) Morningness to depression (diagnosis and treatment). c) Depression to morningness. IVW, inverse-variance weighting; WME, weighted median estimator; 95% CI, 95% confidence interval. Odds ratio: odds of having the outcome for 1-log-odd of increase in the exposure.

#### Associations between sleep duration and depression

No associations were found between genetically predicted sleep duration and diagnosis of depression (OR=0.97, 95%CI=0.80; 1.18) (Figure 2a, Supplementary Tables 4 & 7) or diagnosis or treatment of depression (OR=0.94, 95%CI=0.80; 1.10) (Figure 2b, Supplementary Tables 4 & 7). Results showed an association between genetically predicted depression and shorter sleep duration only with the WME (β=-4.67 minutes, 95%CI=-7.44; -1.89) (Figure 2c, Supplementary Tables 4 & 7).

**Figure 2.**
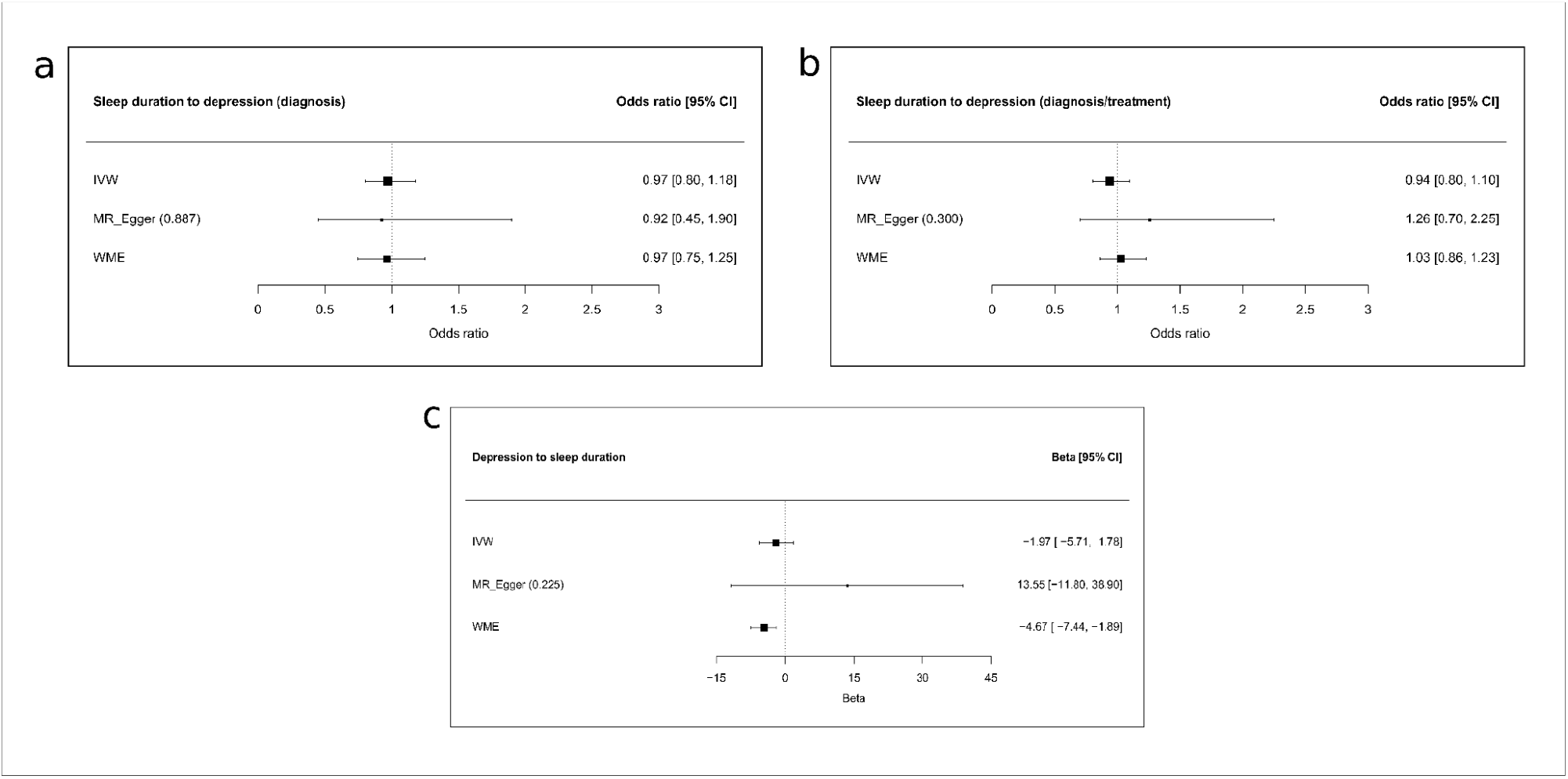
Associations between sleep duration and depression, including sensitivity analyses. a) Sleep duration to depression (diagnosis). b) Sleep duration to depression (diagnosis and treatment). c) Depression to sleep duration. IVW, inverse-variance weighting; WME, weighted median estimator; 95% CI, 95% confidence interval. Odds ratio: odds of having the outcome for 1-log-odd of increase in the exposure. Beta (unstandardised beta coefficient): differences in the outcome for every 1-unit increase in the exposure.

#### Associations between insomnia symptoms and depression

No associations were found between genetically predicted any insomnia symptoms and diagnosis of depression (OR=0.94, 95%CI=0.80; 1.10) or diagnosis or treatment of depression (OR=0.96, 95%CI=0.83; 1.10) (Figure 3a & 3b, Supplementary Tables 5 & 7). Genetically predicted depression was associated with higher odds of having any insomnia symptoms (OR=1.22, 95%CI=1.11; 1.34). MR-Egger indicated no unbalanced horizontal pleiotropy (MR-Egger intercept P-values > 0.05). The MR-Egger slope was not directionally consistent with the IVW estimate. However, the WME estimate was consistent in terms of direction and size (Figure 3c, Supplementary Tables 5 & 7).

**Figure 3.**
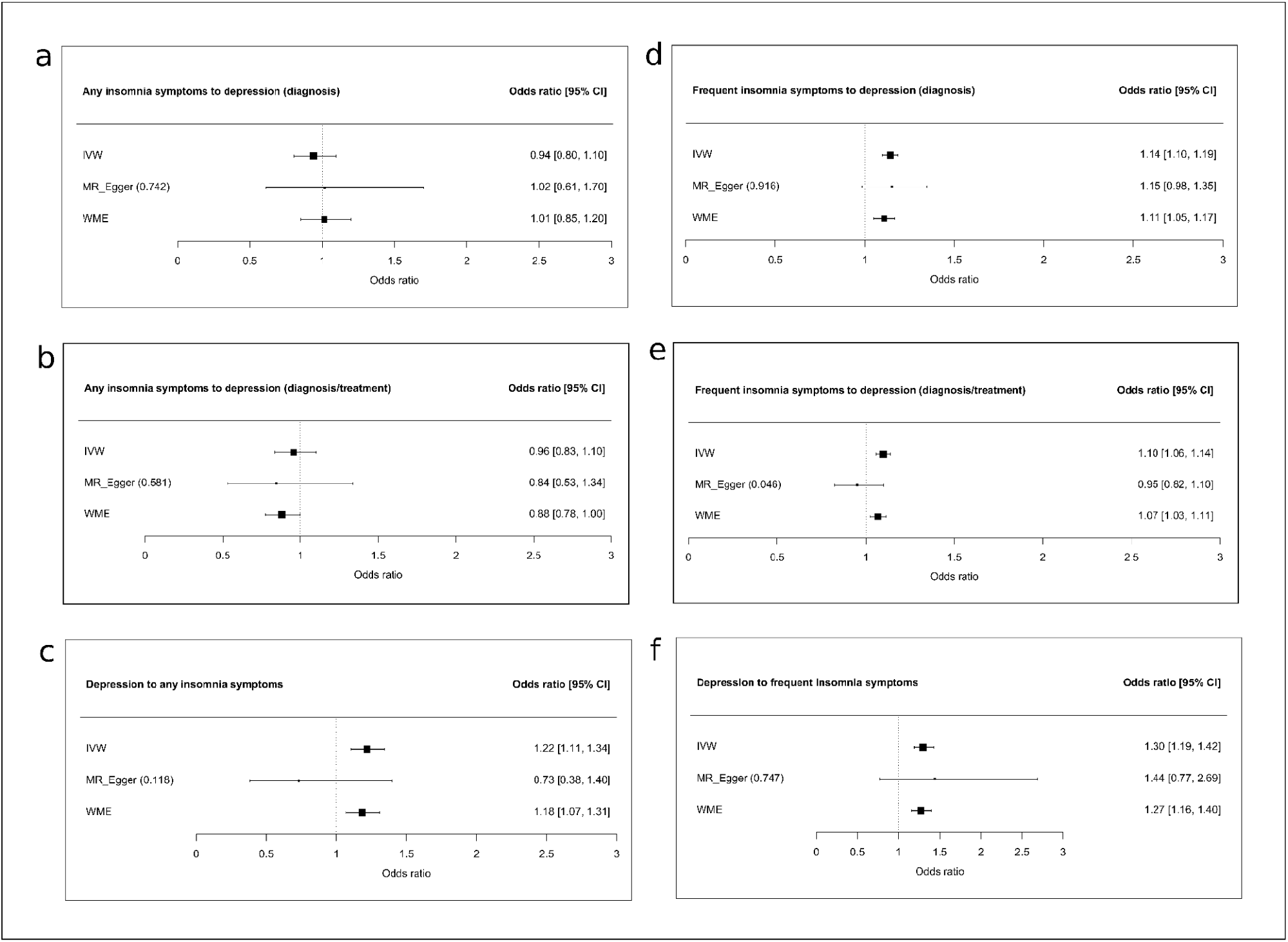
Associations between insomnia and depression, including sensitivity analyses. a) Any insomnia symptoms to depression (diagnosis). b) Any insomnia symptoms to depression (diagnosis and treatment). c) Depression to any insomnia symptoms. d) Frequent insomnia symptoms to depression (diagnosis). e) Frequent insomnia symptoms to depression (diagnosis and treatment). f) Depression to frequent insomnia symptoms. IVW, inverse-variance weighting; WME, weighted median estimator; 95% CI, 95% confidence interval. Odds ratio: odds of having the outcome for 1-log-odd of increase in the exposure.

Genetically predicted frequent insomnia symptoms were associated with higher odds of having a diagnosis of depression (OR=1.14, 95%CI=1.10; 1.19) (Figure 3d, Supplementary Tables 6 & 7) and diagnosis or treatment of depression (OR=1.10, 95%CI=1.06; 1.14) (Figure 3e, Supplementary Tables 6 & 7), and genetically predicted depression was associated with higher odds of having frequent insomnia symptoms (OR=1.30, 95%CI=1.19; 1.42) (Figure 3f, Supplementary Tables 6 & 7). Both MR-Egger and WME approaches indicated no unbalanced horizontal pleiotropy (MR-Egger intercept P-values ≥ 0.05) and were consistent in terms of direction and size (although the “diagnosis or treatment” result from the MR-Egger was not directionally consistent).

### Additional MR results

#### Associations between morningness, sleep duration, insomnia and anxiety outcomes (mixed anxiety-depressive symptoms and anxiety disorders)

No associations were found between genetically predicted morningness (OR=0.97, 95%CI=0.91; 1.03) or genetically predicted sleep duration (OR=0.87, 95%CI=0.75; 1.00) and mixed anxiety-depressive symptoms (Figure 4a & Figure 4b, Supplementary Table 3-6). Results showed that genetically predicted any insomnia symptoms were associated with higher odds of having mixed anxiety-depressive symptoms (OR=1.19, 95%CI=1.08; 1.32). Both MR-Egger and WME approaches indicated no unbalanced horizontal pleiotropy (MR-Egger intercept P-values > 0.05). The MR-Egger slope was not directionally consistent with the IVW estimate, but the WME estimate was (Figure 4c, Supplementary Table 3-6). Genetically predicted frequent insomnia symptoms were associated with higher odds of having mixed anxiety-depressive symptoms (OR=1.20, 95%CI=1.17; 1.23). The MR-Egger approach was consistent in terms of direction. However, the intercept p-value was 0.007; thus, we performed a leave-one-out analysis [60]. The analysis did not suggest undue influence by single outliers or systemic bias (Supplementary Table 8). WME estimate was consistent in terms of direction and size (Figure 4d, Supplementary Table 3-6). No associations were found between genetically predicted morningness (OR=0.92, 95%CI=0.78; 1.09) (Figure 4e, Supplementary Table 3-6), sleep duration (OR=1.09, 95%CI=0.80; 1.47) (Figure 4f, Supplementary Table 3-6), any insomnia symptoms (OR=1.03, 95%CI=0.82; 1.29) (Figure 4g, Supplementary Table 3-6) or frequent insomnia symptoms (OR=1.02, 95%CI=0.95; 1.10) (Figure 4h, Supplementary Table 3-6) with anxiety disorders.

**Figure 4.**
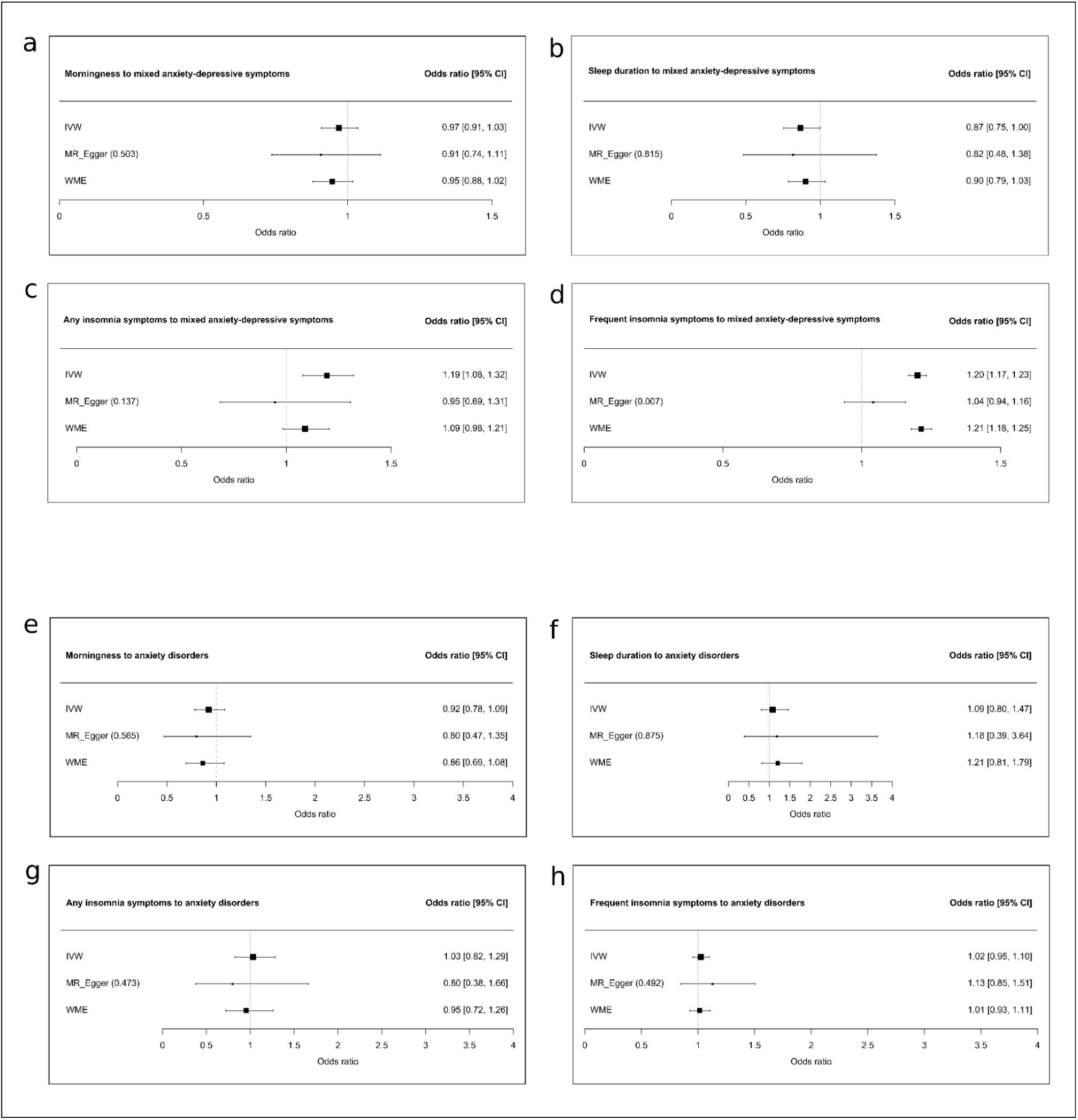
Associations between any insomnia, frequent insomnia, morningness, sleep duration and anxiety outcomes, including sensitivity analyses. a) Morningness to mixed anxiety-depressive symptoms. b) Sleep duration to mixed anxiety-depressive symptoms. c) Any insomnia symptoms to mixed anxiety-depressive symptoms. d) Frequent insomnia symptoms to mixed anxiety-depressive symptoms. e) Morningness to anxiety disorders. f) Sleep duration to anxiety disorders. g) Any insomnia symptoms to anxiety disorders. h) Frequent insomnia symptoms to anxiety disorders. IVW, inverse-variance weighting; WME, weighted median estimator; 95% CI, 95% confidence interval. Odds ratio: odds of having the outcome for 1-log-odd of increase in the exposure.

### Testing MR Assumption III

After Bonferroni corrections, we observed that several genetic variants were associated with some covariates (especially with education and BMI) (Supplementary Table 9).

## Discussion

The present study investigated the association between several circadian and sleep phenotypes and depression using a comprehensive Mendelian randomisation design. We found a causal bidirectional association between depression and insomnia, and suggestive evidence of an effect of morningness on lower odds of depression, and of depression on shorter sleep duration. We also demonstrated that insomnia confers an effect on mixed anxiety-depressive symptoms, while no associations were found between the circadian and sleep traits studied and anxiety disorders. Contrary to the predominant view of circadian rhythm disruptions only as a secondary consequence of depression, this study follows current evidence suggesting a more influential, causal role of these traits on this condition [18].

Our main finding demonstrated bidirectional associations between depression and insomnia following previous cohort [11,61] and MR studies [29–33]. Notably, we found that frequent insomnia, but not any insomnia symptoms, confers an effect on depression. Nowadays, it is widely accepted that insomnia is a risk factor for the development of depression [11], as confirmed by meta-analyses of longitudinal studies [62,63]. In the UKB, a recent machine learning analysis found that one of the best predictors of depression are insomnia symptoms [64]. However, despite advances in the field, the pathway from insomnia to depression is still unclear. Riemann et al. (2012) [65] point towards rapid eye movement (REM) sleep instability. The authors suggest that REM fragmentation in individuals with insomnia impedes correct overnight emotional regulation processes and, when it becomes frequent, heightens responses to negative stimuli and facilitates the development of depression. This theory is supported by experimental studies showing that induced sleep deprivation alters the processing of emotional information [66,67]. Other studies have proposed that insomnia and sleep deprivation may lead to dysregulations in the stress and inflammatory systems, brain neurotrophic factors and monoaminergic, glutamatergic and orexinergic neurotransmission, contributing to mood dysregulations [68].

We also found that depression confers an effect on any and frequent insomnia symptoms. Insomnia has long been recognised as a vegetative sign of depression [10]. Previous observational [16,61] and MR studies [29–33] have found that depression increases the risk of subsequent insomnia, which is a common complaint in this population [69]. Notably, in contrast to some previous MR studies [29,30], we found that depression exerts a larger effect on insomnia than vice-versa. Interestingly, research has revealed shared genetic and neurobiological mechanisms among these conditions [29,30,70], leading to the proposal that the aetiology of insomnia is even more closely associated with biological processes found in psychiatric disorders, such as depression, rather than those involved in regulating sleep-wake cycles [71].

Our results also indicate suggestive evidence of an association between morningness and lower odds of depression but no evidence of an effect of genetically predicted depression on morningness. Numerous cross-sectional studies have reported that morning types are less prone to depressive symptoms, diagnosed depression and antidepressant use than evening types [12]. Accordingly, a randomised clinical trial demonstrated that shifting sleep timing to early hours in evening types leads to lower depressive symptoms [72]. Further, Jones et al. (2019) [42] reported a negative genetic correlation between morningness and diagnosis of depression. However, the directionality of this association is still unclear [73]. Some MR studies have found robust [21,22] and suggestive evidence [33,42] that morningness decreases the risk of depression. However, other MR studies have not found causal associations between morningness and depression [23,35]. A recent study has suggested causality from circadian phase disturbances to mood symptoms using transfer entropy, a non-linear causal inference method [74]. Possible mechanisms for the association between diurnal preferences and depression include better alignment with typical work-rest schedules and higher daily light exposure [75,76]. Moreover, a recent cross-sectional analysis and genome-wide association study have proposed that sleep inertia drives the association of evening types with psychiatric disorders such as depression [77]. Our findings support the well-documented association between eveningness and depression [12–14], and suggest that, at least in the context of modern social schedules, being a morning type may be more beneficial for mental health.

A suggestive effect of depression on shorter sleep duration was also found, while no associations were found between genetically predicted sleep duration and depression. In line with our results, Sun et al. (2022) [33] observed a suggestive inverse association of depression with sleep duration using MR. Moreover, a three-year prospective study found that older adults with incident depression were more likely to shift from an adequate sleep duration to shorter sleep duration [78]. Additionally, both genetic [34] and non-genetic [58] studies have revealed associations between sleep duration and depression. A more thorough examination of this association is warranted since prior studies have identified U-shaped associations between sleep duration and depression [34,79], which could not be assessed in this work due to the potential for biased results from nonlinear MR methods [80].

Additional results showed that any and frequent insomnia symptoms were associated with higher odds of having mixed anxiety-depressive symptoms, while no associations were found with the other circadian and sleep phenotypes studied. These results support our main findings; insomnia is the trait more strongly associated with depression (even when it is measured mixed with anxiety symptoms). Notably, both any and frequent insomnia symptoms exert an influence on mixed anxiety-depressive symptoms, while only frequent symptoms confer an effect on depression. We speculate that this could be attributed to the outcome being more nuanced, allowing for the effect to be observed even with mild insomnia symptoms. Moreover, we did not find evidence of an effect of circadian and sleep traits on anxiety disorders, implying a unique association with depression that is not replicated when the outcome includes only anxiety. There is considerable literature documenting circadian and sleep problems in individuals with anxiety disorders [81–83]. However, most of these studies were observational in nature, precluding causal inference. In the case of insomnia, a meta-analysis of longitudinal studies indicated that it increased the risk of anxiety disorders [62], which is consistent with a recent MR study [84]. We did not observe this association, which may be due to discrepancies in how anxiety was measured. The lack of association between morningness and anxiety follows previous literature, including MR studies [12,85], as well as the association between sleep duration and anxiety disorders [84]. Considering that anxiety still lacks robust instruments to perform MR analyses, future studies should assess its effect on circadian and sleep traits.

Despite substantial observational evidence linking depression with circadian and sleep traits [12–15,17,18], consistent causal associations have been difficult to establish. It is thus noteworthy that we found bidirectional causal associations between depression and insomnia symptoms. These associations are robust even though they rely on an estimation of insomnia symptoms based solely on one self-reported question, and they are specific as they were confirmed for insomnia symptoms but not for morningness or sleep duration. Therefore, it is important to note that a persistent challenge in the chronobiological field is the distinction between traits; for example, early morning awakening, a classic feature of insomnia [86] (as well as depression [87]), might be misinterpreted as a morning preference. Future research should explore whether distinct components of insomnia are causally related to depression and performing in-depth explorations into the mechanisms driving these associations.

Finally, the three core MR assumptions were corroborated to validate our findings. We instrumented the best genetic variants available (assumption I), implemented sensitivity analyses to check horizontal pleiotropy (assumption II), and performed regressions between our genetic instruments and unobserved confounders (assumption III). Regarding this last assumption, we found that some of the variants instrumented were associated with common confounders. These associations warrant further examination, as they could be related to the traits being polygenic in nature and/or may constitute vertical rather than horizontal pleiotropy. Future research shall explore whether any of the SNPs are vertically pleiotropic by conducting MR mediation analyses [88].

## Strengths and limitations

The study’s strengths include using an MR approach, which reduces bias due to confounding and reverse causality, a large sample size that ensures sufficient power, carefully selecting genetic instruments [45], and assessing several circadian, sleep and depression phenotypes. However, limitations should also be acknowledged. First, given that UKB participants are healthier than the general population, the findings may not apply to individuals with significant comorbidities [89]. Second, the sample was restricted to white Europeans, as the genetic variants were discovered in this population; future work should examine these genetic associations in diverse racial and ethnic groups for generalisation and to promote health equity. Third, volunteers were 40-69 years old; thus, our results cannot be generalised to other age groups. Fourth, most of our exposures were binary, which has some limitations [90]. Fifth, the circadian and sleep phenotypes were assessed using self-reported questions, emphasising the need for replication using objective measures such as accelerometer-derived data when possible. Sixth, we found that several genetic variants were linked to confounders, and we cannot rule out the possibility of other confounding factors. Last, depression is a highly heterogeneous condition [91,92], underscoring the need to examine its subtypes (e.g., typical and atypical; mild, moderate and severe; episodic, recurrent and persistent).

## Conclusions

In conclusion, this MR study offers novel evidence of causal associations between circadian rhythms, sleep, and depression through the analysis of multiple phenotypes. Notably, the findings demonstrate bidirectional causal associations between depression and insomnia, as well as suggestive evidence linking morningness with lower odds of depression and an effect of depression on sleep deprivation. These findings provide insights into the association between the circadian system and depression, enhancing our understanding of the etiopathogenic processes involved in this condition. Future studies should focus on depressive subtypes and other circadian and sleep traits, and replicate these findings using other datasets and methods.

## Data Availability

The UK Biobank data are publicly available to all bona fide researchers at https://www.ukbiobank.ac.uk.

## Acknowledgements

This research has been conducted using the UK Biobank Resource under Application Number 68232. This work uses data provided by patients and collected by the NHS as part of their care and support. We thank all UKB researchers and volunteers. We thank Albert Henry for his suggestions on analyses. VP was supported by Programa de Desarrollo de las Ciencias Básicas (PEDECIBA, MEC-UdelaR, Uruguay), Agencia Nacional de Investigación e Innovación (ANII, Uruguay) [grant number MOV_CA_2020_1_163153], Comisión Sectorial de Investigación Científica (CSIC, UdelaR, Uruguay) and, Comisión Académica de Posgrados (CAP, UdelaR, Uruguay). MR is funded by the UK Medical Research Council (grants MC_UU_00019/1 and 3). BT and AS were supported by Comisión Sectorial de Investigación Científica (CSIC, UdelaR, Uruguay) [grant number 883158]. VG was supported by the Professor David Matthews Non-Clinical Fellowship from the Diabetes Research and Wellness Foundation (United Kingdom) [grant number SCA/01/NCF/22].

## Ethics approval

UKB participants had given written informed consent and ethical approval for the study was granted by the North West Haydock Research Ethics Committee of the UK’s Health Research Authority.

